# Excess hypoglycaemia-related hospitalisation in older adults with comorbid type 2 diabetes and dementia: a UK Biobank cohort study

**DOI:** 10.64898/2026.09.15.26363121

**Authors:** Busra Donat-Ergin, Ben Au-Yeung, Katharina Mattishent, Mizanur Khondoker, Anne Marie Minihane, Richard Ian Gregory Holt, Jay Amin, Ketan Dhatariya, Michael Hornberger

## Abstract

**Background:** Older adults living with both type 2 diabetes mellitus (T2DM) and dementia may be especially vulnerable to severe hypoglycaemia because cognitive impairment complicates glucose monitoring, medication management and recognition of glycaemic symptoms. It remains unclear whether the combined conditions confer an excess risk of hypoglycaemia-related hospitalisation beyond the risks associated with either condition alone.

**Aim:** To determine whether comorbid T2DM and dementia are associated with excess hypoglycaemia-related hospitalisation. Secondary aims were to characterise overall hospital admissions, falls-related admissions, hyperglycaemia-related admissions and mortality across disease states.

**Methods:** We analysed retrospective longitudinal UK Biobank data for 329,566 participants aged 65 years or older. T2DM and dementia were modelled as time-varying exposures. Recurrent hospital admissions were analysed using Prentice-Williams-Peterson total-time models and mortality using a time-dependent Cox model, adjusted for sex and socioeconomic deprivation. We reported joint effects, condition-specific effects and interaction on multiplicative and additive scales; additive interaction was quantified using the relative excess risk due to interaction (RERI).

**Results:** By the end of follow-up, 18,397 participants (5.6%) had developed T2DM only, 6,214 (1.9%) dementia only and 802 (0.2%) both conditions. Hypoglycaemia-related hospitalisation occurred in 6.61% of participants while living with both conditions, compared with 1.88% with T2DM only, 0.91% with dementia only and 0.15% with neither condition. Relative to neither condition, the adjusted hazard ratio (HR) was 21.28 (95% CI 17.76 to 25.50) for T2DM only, 14.73 (10.47 to 20.71) for dementia only and 104.94 (73.79 to 149.24) for comorbidity. T2DM increased the hypoglycaemia admission hazard among people with dementia (HR 7.13, 4.63 to 10.97), while dementia increased it among people with T2DM (HR 4.93, 3.60 to 6.76). The positive additive interaction was substantial (RERI 69.93, 35.17 to 104.68; p<0.001), although interaction was sub-multiplicative (HR 0.33, 0.21 to 0.53). Comorbidity was also associated with the highest overall admission, falls and mortality burden, but these outcomes did not show clear positive additive interactions.

**Conclusion:** Hypoglycaemia-related hospitalisation was the principal excess comorbid outcome in older adults with T2DM and dementia. The combined hazard substantially exceeded that expected from adding the separate relative hazards associated with each condition. General admissions, falls and mortality were also elevated in the comorbid state, corroborating existing evidence and indicating wider clinical vulnerability. These findings support prioritising safer, cognitively appropriate glycaemic management in this high-risk population.

## Introduction

Type 2 diabetes mellitus (T2DM) and dementia are both highly prevalent conditions [1], [2] and represent major public health challenges in ageing societies [3]. The global burden of T2DM continues to rise, driven by demographic ageing, health behaviours, and increasing obesity rates [1], [4]. Similarly, dementia prevalence is steadily increasing, with substantial implications for individuals, carers, and health systems. As both conditions are common in older adults, it is unsurprising that their co occurrence is frequent, leading to a growing population of individuals living with comorbid T2DM and dementia [5]. Existing research indicates that diabetes is one of the most frequent comorbidities observed in individuals with dementia, particularly among those aged over 65 years, reflecting the high prevalence of both diseases in older age groups [3], [6], [7], [8].

Biological and clinical mechanisms underpinning the relationship between T2DM and dementia are increasingly understood to be bidirectional. Diabetes contributes to neurodegeneration through chronic hyperglycaemia, insulin resistance, microvascular pathology, and systemic inflammation, all of which have been implicated in the development of both Alzheimer’s disease and vascular dementia [7]. Conversely, dementia related cognitive impairment reduces the capacity for safe diabetes self management, including medication administration, glucose monitoring, meal planning and recognition or communication of glycaemic symptoms [9]. This combination makes severe hypoglycaemia a particular concern. Hypoglycaemia is an established cause of emergency hospitalisation in people with diabetes [10], while dementia may increase vulnerability through impaired self-management and dependence on carers. Severe hypoglycaemic events have also been associated with subsequent dementia risk [11], [12], [13], suggesting a bidirectional relationship.

Although older adults with either T2DM or dementia experience substantial hospital use, previous research has largely described overall admission burden or compared static diagnostic groups [14]. By contrast, relatively little is known about how comorbid T2DM and dementia affects hospital admission rates, risk factors and mortality. Such comparisons do not establish whether the comorbid T2DM and dementia produces more harm than would be expected from the separate effects of each condition. This distinction is clinically important for hypoglycaemia, as an excess joint effect would identify people with both conditions as a priority group for prevention, treatment review and supported glucose management. Current evidence lacks in this regard.

The primary aim of this study was, therefore, to determine whether comorbid T2DM and dementia are associated with excess hypoglycaemia-related hospitalisation beyond the relative hazards associated with either condition alone. Secondary aims were to characterise overall hospital admissions, falls-related and hyperglycaemia-related admissions, and mortality across: T2DM only, dementia only, both conditions, and neither condition. We hypothesised that hypoglycaemia-related admissions would show positive additive interaction between T2DM and dementia. The other outcomes were examined to establish the broader hospital and mortality context of this comorbidity.

## Methodology

### Study Design and Population

The current study used retrospective longitudinal data from the UK Biobank. At baseline 503,317 40- to 69-year-olds were recruited between 2006 and 2010 across England, Scotland and Wales. They answered questionnaires providing their demographical information and gave consent for their NHS data to be used. Participants who were 65 years old and over were eligible for the study. People with T1DM were excluded from the study. Overall, the final sample contained 329,566 participants.

### Ethics

Ethical approval for the UK Biobank was granted by the North West - Haydock Research Ethics Committee (reference no. 21/NW/0157), and all participants provided written informed consent.

### Outcome Variables

Hospital admissions data were obtained from the Hospital Episode Statistics - Inpatient (HESIN) data set. Under HESIN, admissions are broken into episodes. Some episodes from the same admission were sometimes recorded separately, so we combined overlapping episodes. Only in-patient admissions that admissions that involved at least one overnight stay were included in the analysis. Mortality data came from national death registries or relatives’ reports. With these data, we measured participants’ admission risk factors and mortality hazards.

### Covariates

Covariates were taken at baseline using the online questionnaires. Reasons for hospital admission were determined by examining ICD-10 diagnostic codes across participants’ admission episodes [15]. Fall-related admissions were identified using codes W00–W19. Hypoglycaemia admissions were defined using codes E15, E16.0, E16.1, E16.2. Hyperglycaemia admissions were identified using codes E10.1, E11.1, E13.1, E14.1 (associated with diabetic ketoacidosis) and R73.9.

Sex (male vs female) and deprivation quintiles were the two co-variates. In the UK, deprivation is measured using the index of multiple deprivation (IMD) which scores residential areas on factors such as education, crime, employment and health. Each nation (England, Scotland and Wales) uses varying factors and weights, so scores are ranked relative to their nation. These ranks produce deprivation quintiles where 1 is the least deprived and 5 is the most deprived. Finally, we attempted to take ethnicity (White/Asian/Black/ Chinese/ Mixed) into account but there were not enough non-white participants for comparison.

### Statistical Analysis

A time-dependent Cox proportional hazards model was used to estimate mortality hazards. The first risk interval began at age 65, unless the participant was admitted to hospital during their 65th birthday, in which case follow-up started at discharge. Starting from the 65th birthday ensures that only geriatric admissions are used. Subsequent risk intervals were created at changes in disease status (T2DM or dementia diagnosis). Follow-up ended at death, leaving the UK, or November 2022 where hospital records ended. The beginning and end of each interval was measured in years from the participant’s 65^th^ birthday.

Prentice Williams Peterson total time (PWP-TT) models were then used to assess risk factors of hospitalisation. Creation of risk intervals can be seen in Figure 1. Time began at 65 years old and stayed continuous throughout follow up. Additional risk intervals were created at each hospital admission with the following interval starting at discharge. This creates a gap as participant cannot be at risk while already admitted. Therefore, if disease onset occurred during a hospital admission, the disease change was applied post discharge, rather than creating a new interval during admission. PWP-TT models were chosen over PWP gap time models as we were interested in cumulative admission risks instead of speed of admission post-discharge. PWP-TT was also used to assess admission risks for falls, hypoglycaemia and hyperglycaemia. All models were stratified by admission number. High frequency re-admissions were sparse, so general admissions were truncated at 10 admissions, and admissions for falls, hyperglycaemia and hypoglycaemia were capped at two. Fewer than 1% of participants exceeded these truncation thresholds. Participants were censored after truncation.

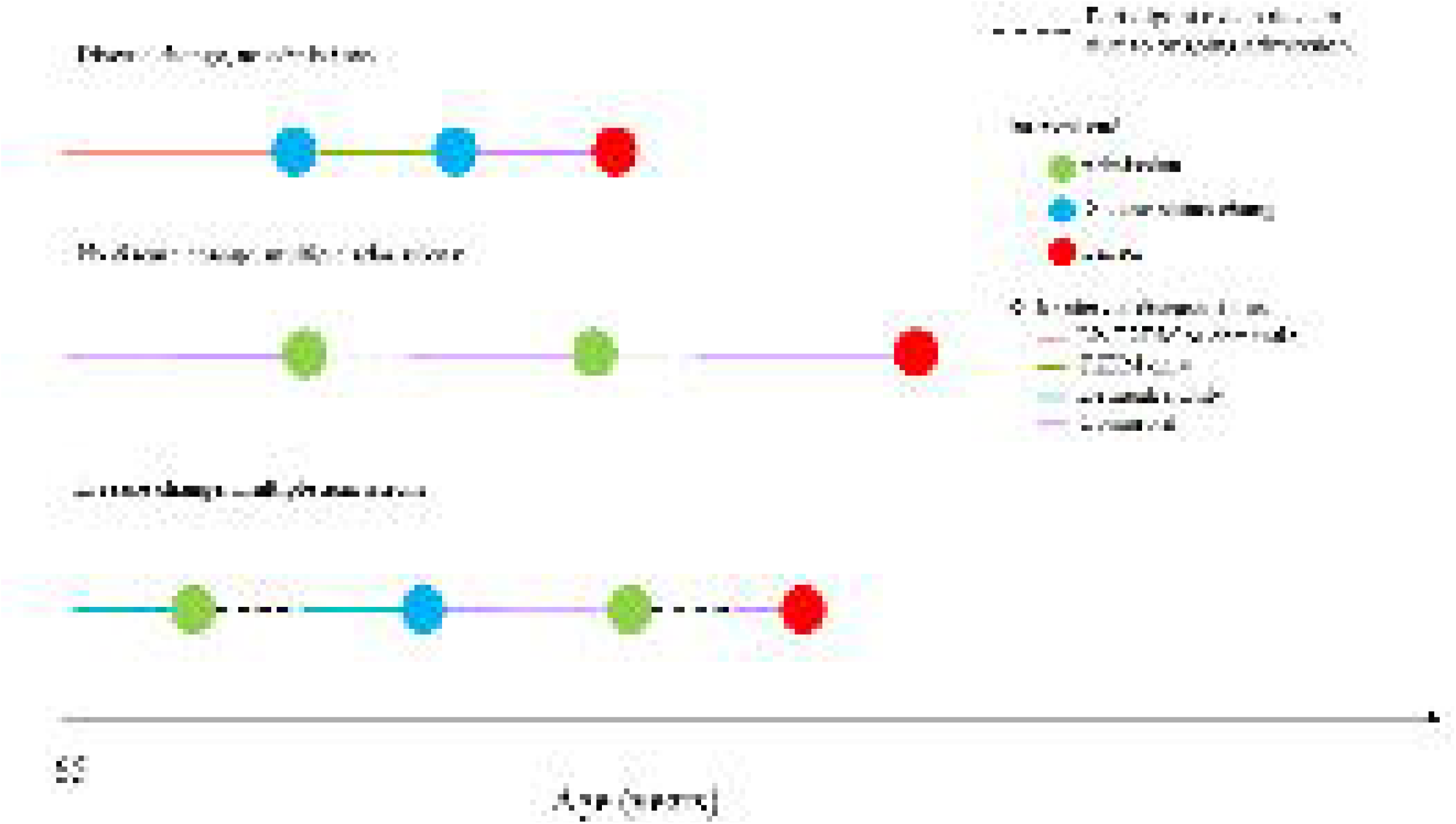

All the models were run with a dementia x T2DM main effects and interaction with sex and deprivation as co variates. Dementia and T2DM status were time-varying defined according to whether the participant was diagnosed at the start of the risk interval. Sex and deprivation were static variables taken at screening. The models were clustered by participant ID to account for within participant correlation. To make the effects of dementia and T2DM on the outcomes clearer we follow a recommended report template [16]. First the joint effects of dementia and T2DM are presented with living without diabetes or dementia as the reference. The effects of dementia conditional on T2DM status, and vice versa, are then presented to illustrate effect modification. Finally, interaction is reported on the multiplicative scale (the models’ natural scale) and on the additive scale using the relative excess risk due to interaction (RERI). Positive multiplicative and additive interactions indicate an excessive comorbid hazard, greater than the produce/ sum of the individual dementia and T2DM effects respectively. Conversely, a sub-multiplicative or additive interaction would indicate that comorbidity does not carry excessive risks as one disease attenuates the effect of the other. To account for multiple testing of 5 models, we used a Bonferroni correction, setting alpha at 0.01.

To calculate the probability of experiencing an admission for each disease status (no T2DM or dementia, T2DM-only, dementia-only and comorbid), we divided the number of participants who experienced at least one admission by the total number of participants for each disease status. Admission rates were calculated for each disease-status by dividing the total number of admissions by the total years participants spent at that disease status.

## Results

### Cohort Characteristics

Our final sample had 329,566 participants. By the end of the study 304,153 (92.3%) developed neither T2DM nor dementia, 18,397 (5.6%) had T2DM-only, 6,214 (1.9%) had dementia-only, and 802 (0.2%) developed both T2DM and dementia. Table 1 shows transition in disease status from study entry to prior to censor. Table 2 shows their characteristics. There were no sex differences between those who reached each disease status, χ² (3) = 4.17, p = .244, V = .10. Small, but significant differences in deprivation were found, *F* (3, 3127) = 556.0, *p* < .001, η² = .005. Deprivation was highest among those who developed T2DM-only or comorbid T2DM and dementia (2.8 ± 1.5 and 2.9 ± 1.5, respectively), with no significant difference between the two (*p* = 0.02). Deprivation was lower among those who developed dementia-only (2.6 ± 1.4) and lowest among those who did not develop T2DM or dementia (2.4 ± 1.3), with small effect sizes across comparisons (*d* = 0.14–0.39).

### Hypoglycaemia-related admissions

Hypoglycaemia-related hospitalisation occurred in 6.61% of participants with comorbid T2DM and dementia, compared with 1.88% T2DM only, 0.91% dementia only and 0.15% neither condition. The admission rate was 0.04 per person-year in the comorbid state and below 0.01 per person-year for the other conditions (Table 3). Relative to neither condition, the adjusted risk was 21.28 (95% CI 17.76-25.50) for T2DM only, 14.73 (10.47-20.71) for dementia only and 104.94 (73.79-149.24) for comorbid T2DM-dementia (Table 4). Among participants living with dementia, the presence of T2DM increased the risk of hypoglycaemia-related admission 7.13-fold (95% CI 4.63-10.97). Among participants living with T2DM, the presence of dementia increased the risk 4.93-fold (95% CI 3.60-6.76). The joint effect showed strong positive additive interaction (RERI 69.93, 95% CI 35.17 to 104.68; p<0.001), although this was sub-multiplicative (HR 0.33, 95% CI 0.21 to 0.53; p<0.001), indicating excess hypoglycaemia-related admission risk.

### General hospital admissions

The probability of being hospitalised at least once was highest when living with comorbid T2DM-dementia (87.41 %), followed by dementia-only (84.86%), then T2DM-only (60.88%), and finally controls (43.94%). Corresponding admission rates were 1.08, 0.81, 0.32 and 0.12 admissions per person-year (Table 3, Figure 2). Relative to controls, risks were higher for T2DM only (HR 1.28, 95% CI 1.26-1.30), dementia only (HR 1.89, 1.82-1.95) and both conditions (HR 1.90, 1.75-2.06). Within each condition, dementia increased admission risks for people with T2DM (HR 1.48, 1.36-1.61), whereas T2DM did not significantly increase risks for people with dementia (HR 1.01, 0.92-1.10). The interaction for this effect was sub-multiplicative (HR 0.79, 0.72-0.86). The RERI estimate was negative (−0.27, −0.44 to −0.10), indicating that the comorbid risk did not exceed the sum of the risk for each condition.

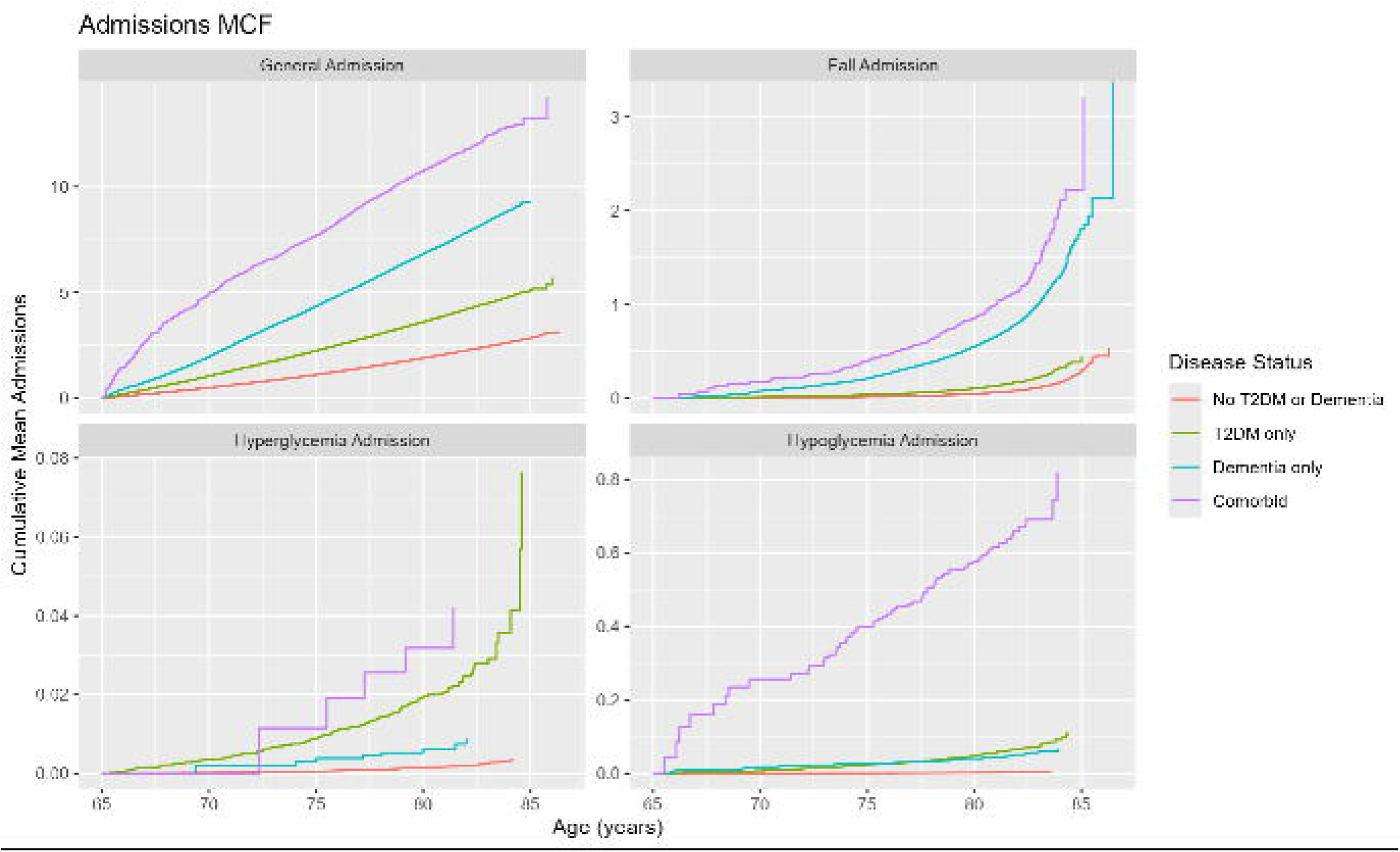

### Falls-related admissions

Falls-related admissions occurred in 17.71% comorbid T2DM-dementia, 14.82% dementia-only, 4.28% T2DM-only and 2.41% controls. Relative to controls, the risk was 9.08 (95% CI 7.57-10.90) for both conditions, 6.66 (6.14-7.22) for dementia only and 2.02 (1.87 to 2.19) for T2DM only. Within each condition, dementia increased the falls admission risk for T2DM (HR 4.49, 3.69-5.45), while T2DM showed a small significant increase for dementia (HR 1.36, 1.13-1.65). The interaction was sub-multiplicative (HR 0.67, 0.55-0.83), and the RERI confidence interval included zero (RERI 1.40, −0.29-3.09), providing no clear evidence of positive additive interaction for this risk factor.

### Hyperglycaemia-related admissions

Hyperglycaemia-related admissions were rare, occurring in 0.75% comorbid participants, 0.72% T2DM-only, 0.16% dementia-only; and 0.09% controls. Relative to controls, the risk was 15.94 (95% CI 6.47-39.31) comorbid T2DM-dementia, 11.73 (9.17-15.00) for T2DM-only and 3.39 (1.68-6.83) for dementia-only. Within each condition, T2DM increased risks for dementia (HR 4.70, 1.55-14.21), whereas dementia did not significantly increase risks for T2DM (HR 1.36, 0.56-3.31). Neither the multiplicative interaction estimates (HR 0.40, 0.13-1.22) nor the additive interaction estimates (RERI 1.82, −12.43-16.07) provided clear evidence of interaction.

### Mortality

Survival during follow-up was: 93.32% for controls, 85.57% T2DM-only, 52.12% dementia-only, and 49.88% T2DM-dementia. Relative to controls, mortality risks were 2.58 (95% CI 2.47-2.68) T2DM-only, 14.47 (13.90-15.05) dementia-only and 15.99 (14.47 17.66) T2DM-dementia (Table 5; Figure 3). Within each condition, dementia significantly increased mortality for T2DM (HR 6.21, 5.59-6.89), whereas T2DM did not significantly increase mortality for dementia (HR 1.11, 1.00-1.23; p=0.060). The interaction was sub-multiplicative (HR 0.43, 0.38-0.48), and there was no evidence of additive interaction (RERI −0.06, −1.71-1.59).

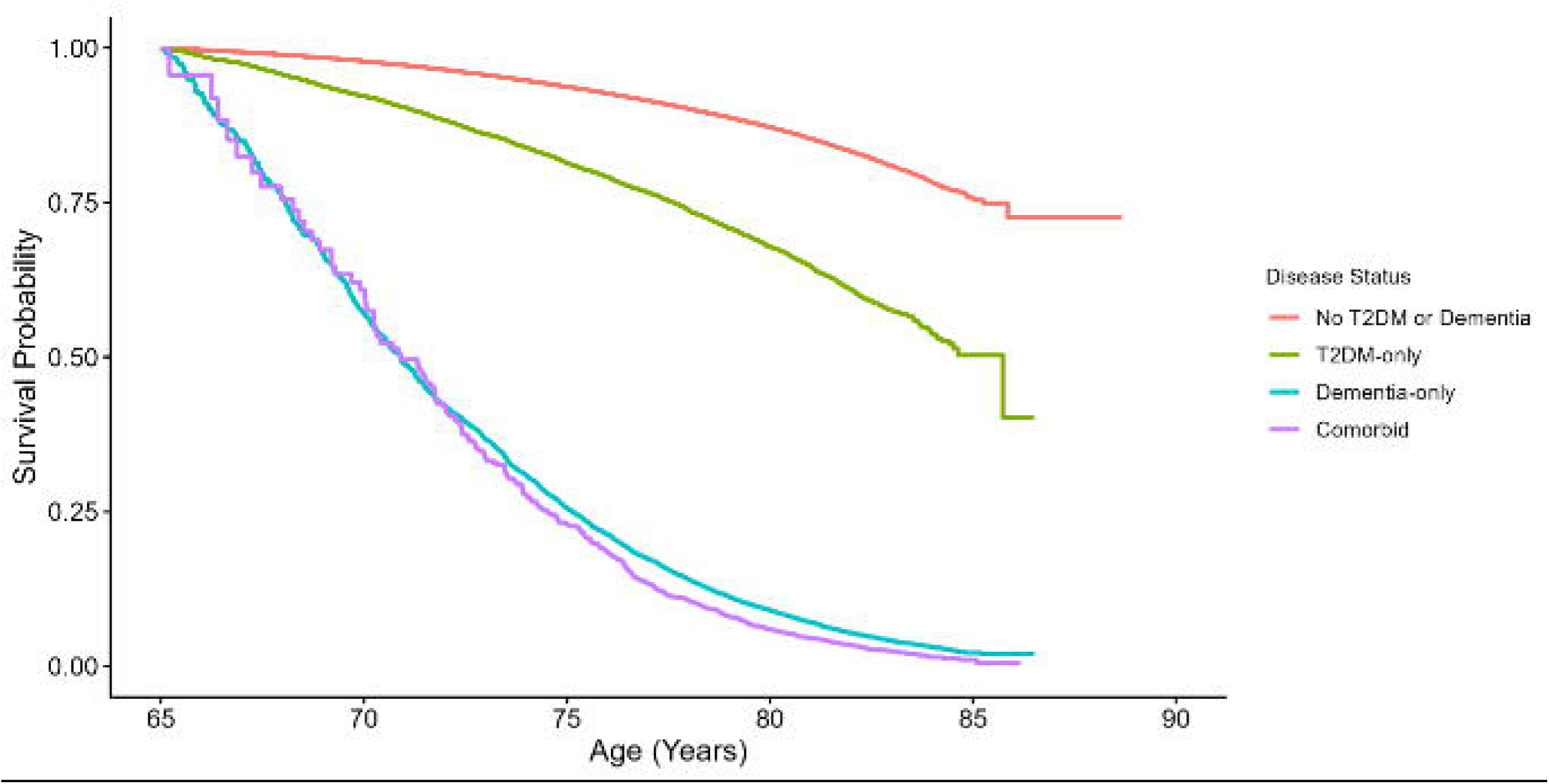

## Discussion

The main finding of our study is that older adults with comorbid T2DM-dementia are at a significantly additive risk for hypoglycaemia-related hospitalisations, compared to each condition alone. Although comorbidity was also associated with the greatest overall burden of hospital admissions, falls and mortality, there was no additive effect for the comorbidity for these risks. By contrast, hypoglycaemia emerges as the distinctive excess comorbid risk with clear clinical implications.

In terms of the hypoglycaemia comorbidity findings, several clinically plausible pathways may contribute to this study. Dementia can impair medication management, meal regularity, glucose monitoring, symptom recognition and the ability to respond appropriately to low glucose [17]. Diabetes treatment may therefore become increasingly difficult to manage safely as cognition declines, particularly when regimens are complex or depend heavily on self-management. Previous studies have reported greater hypoglycaemia risk among people with diabetes and dementia [11], [12], while severe hypoglycaemia has itself been associated with subsequent dementia risk [13].There have been suggestions, that glucose monitoring technology, such as Continuous Glucose Monitoring might improve, glucose management in comorbid patients [18], [19] but the evidence for this remains limit. Our findings show that comorbid T2DM-dementia should be considered a high-risk hypoglycaemia admission group and hence better, future preventative management is needed.

By contrast, the results for the general hospital admission and mortality showed a different pattern. For both risk factors, dementia increased the risk for people with T2DM but not the other way around. It suggests that dementia accounts for much of the higher risk observed in the comorbid condition for general admission and mortality, rather than a combined effect of T2DM and dementia. Previous findings have shown that people with dementia have more frequent hospital admissions than those without dementia [20]. Admission rates and mortality were associated with multimorbidity including diabetes [10], [21], [22], [23], [24], [25]. Our findings extend these findings by demonstrating that dementia drives the admission results, rather than T2DM.

Similarly, falls-related admission showed that, although the comorbidity had overall much higher rates of falls, this effect was mainly due to increased falls because of dementia and not T2DM. There is a well-established association between fall risk and cognitive impairment [26], which likely explains this finding. Specifically, a previous report identified dementia (41.2%) and accidental falls (40.1%) as the leading causes of hospitalisation among older adults [27], further corroborating our findings. Hyperglycaemia-related admissions were low across all groups, and in particular for the comorbid group. Overall, T2DM increased hyperglycaemia-related admission risks for dementia, but the overall prevalence was very low. Interestingly, the hyperglycaemic findings reinforce the specificity of our hypoglycaemia admission finding, since it suggests that the admission risk in the comorbid group is not simply due to dysglycaemic regulation but hypoglycaemia, specifically. Still, future, prospective research is needed to investigate this aspect further.

Out study has several strengths, including a large sample size and longitudinal follow up. However, the main strength lies in the analytical approach, which modelled dementia and T2DM as separate time-varying exposures, allowing participants to change disease state during follow-up. It also enabled estimation of separate, joint and disease-specific associations within a single modelling framework. Further, recurrent hospitalisations were analysed using a PWP total-time approach, and interaction was reported on both additive and multiplicative scales [16]. Overall, this approach allowed a more in-depth investigation of how to common, long-term conditions impact hospital admission rates, risks and mortality.

Several limitations require also consideration. Recorded diagnosis may occur after disease onset, potentially misclassifying some person-time and biasing disease-state estimates. Medication exposure, treatment intensity, glycaemic targets, frailty, functional status and carer involvement were not included, so the mechanisms underlying the excess hypoglycaemia hazard could not be directly examined. The hypoglycaemia outcome captured events requiring overnight hospital admission and therefore does not represent all hypoglycaemic episodes. The comorbid group was relatively small, and hyperglycaemia events were rare, producing wide confidence intervals. The predominantly ethnically white UK Biobank population limited ethnic subgroup analyses and may restrict generalisability. Finally, RERI derived from proportional-hazards models should be interpreted as interaction between relative hazard measures rather than as a direct estimate of an absolute risk difference.

The findings have direct implications for research and service development. Future studies should examine whether medication class, treatment intensity, impaired awareness, nutritional instability, frailty and reduced self-management capacity explain the excess hypoglycaemia hazard. They should also test whether cognitively appropriate treatment review, regimen simplification, individualised glycaemic targets, continuous glucose monitoring or structured caregiver support can reduce severe events. These approaches might allow better prevention of hypoglycaemic admissions in this high-risk group.

In conclusion, comorbid T2DM-dementia was associated with the greatest overall burden of hospitalisation, falls and mortality, but hypoglycaemia-related hospitalisation was the only outcome showing clear positive additive interaction. The coexistence of both conditions, therefore, appears to create a particularly vulnerable clinical group for severe hypoglycaemic events beyond the hypoglycaemic risks associated with either condition alone. Prevention of hypoglycaemia should be a central priority in the care of older adults with comorbid T2DM-dementia.

## Data Availability

All data produced in the present work are contained in the manuscript.

## Appendix Tables

**Table 1:** Transition table.

|  | Final disease status |  |  |  | **Total** |
| --- | --- | --- | --- | --- | --- |
|  | No T2DM or Dementia | T2DM-only | Dementia-only | Comorbid |  |
| Starting disease status |  |  |  |  |  |
| No T2DM or Dementia | 304,153 | 12,498 | 5,936 | 620 | 323,207 (98.1%) |
| T2DM-only | - | 5,899 | - | 136 | 6,035 (1.8%) |
| Dementia-only | - | - | 278 | 22 | 300 (0.1%) |
| Comorbid | - | - | - | 24 | 24 (<0.1%) |
| **Total** | 304,153 (92.3%) | 18,397 (5.6%) | 6,214 (1.9%) | 802 (0.2%) | 329,566 (100.0%) |

**Table 2:** Characteristics by disease status prior to censor.

|  | No T2DM or Dementia<br>(n = 304153) | T2DM-only<br>(n = 18397) | Dementia-only<br>(n = 6214) | Comorbid<br>(n = 802) |
| --- | --- | --- | --- | --- |
| Age at diagnosis, mean ± SD | | | | |
| Dementia | - | - | 75.1 ± 5.5 | 75 ± 5.4 |
| Diabetes | - | 67.9 ± 7.7 | - | 70.8 ± 7.4 |
| Sex, N (%) |  |  |  |  |
| Female | 171277 (56.3%) | 8142 (44.3%) | 3116 (50.1%) | 351 (43.8%) |
| Male | 132876 (43.7%) | 10255 (55.7%) | 3098 (49.9%) | 451 (56.2%) |
| Ethnicity, N (%) |  |  |  |  |
| White | 293538 (96.5%) | 16682 (90.7%) | 6019 (96.9%) | 729 (90.9%) |
| Asian | 3311 (1.1%) | 727 (4.0%) | 50 (0.8%) | 25 (3.1%) |
| Missing | 2832 (0.9%) | 445 (2.4%) | 88 (1.4%) | 22 (2.7%) |
| Other | 1709 (0.6%) | 218 (1.2%) | 20 (0.3%) | 13 (1.6%) |
| Mixed | 1064 (0.3%) | 99 (0.5%) | 20 (0.3%) | 3 (0.4%) |
| Black | 1007 (0.3%) | 162 (0.9%) | 13 (0.2%) | 7 (0.9%) |
| Chinese | 692 (0.2%) | 64 (0.3%) | 4 (0.1%) | 3 (0.4%) |
| Deprivation (1-5), mean ± SD | | | | |
| | 2.4 ± 1.4 | 2.8 ± 1.5 | 2.6 ± 1.4 | 2.9 ± 1.5 |
| Reason censored, N (%) |  |  |  |  |
| Study end | 280562 (92.2%) | 15735 (85.5%) | 3239 (52.1%) | 400 (49.9%) |
| Died | 23358 (7.7%) | 2654 (14.4%) | 2975 (47.9%) | 402 (50.1%) |
| Left the UK | 233 (0.1%) | 8 (<0.1%) | 0 (0) | 0 (0) |

**Table 3:** Admission Probability and Rate.

| Admission | Disease Status | $\geq 1$ Admission Probability (%) | Admission rate (per year) |
| --- | --- | --- | --- |
| General | No T2DM or Dementia | 43.94 | 0.12 |
|  | T2DM-only | 60.88 | 0.32 |
|  | Dementia-only | 84.86 | 0.81 |
|  | Comorbid | 87.41 | 1.08 |
| Falls | No T2DM or Dementia | 2.41 | <0.01 |
|  | T2DM-only | 4.28 | 0.01 |
|  | Dementia-only | 14.82 | 0.07 |
|  | Comorbid | 17.71 | 0.1 |
| Hypoglycaemia | No T2DM or Dementia | 0.15 | <0.01 |
|  | T2DM-only | 1.88 | <0.01 |
|  | Dementia-only | 0.91 | <0.01 |
|  | Comorbid | 6.61 | 0.04 |
| Hyperglycaemia | No T2DM or Dementia | 0.09 | <0.01 |
|  | T2DM-only | 0.72 | <0.01 |
|  | Dementia-only | 0.16 | <0.01 |
|  | Comorbid | 0.75 | <0.01 |

**Table 4:**
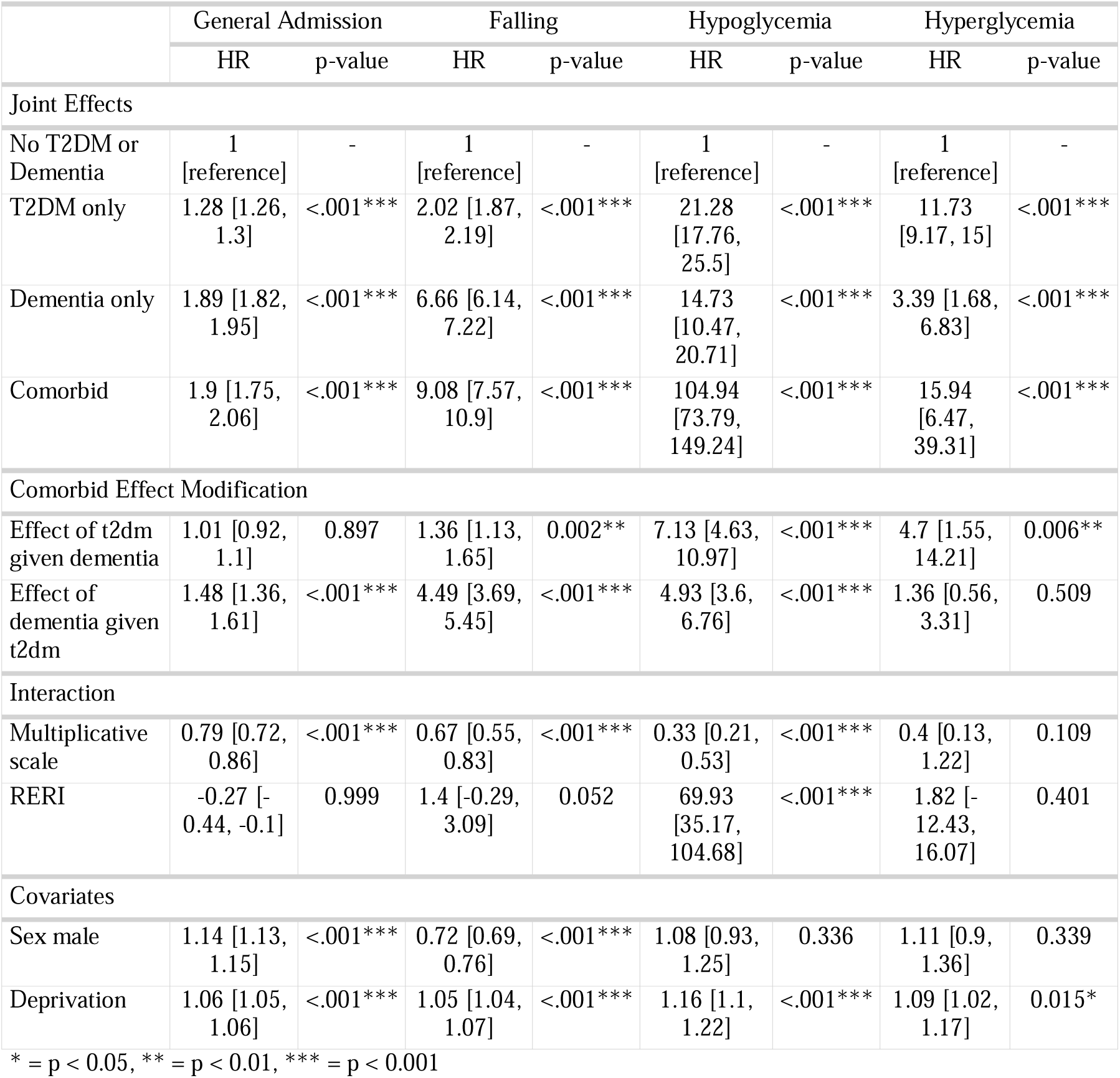
Admission Risk.

|  | General Admission |  | Falling |  | Hypoglycemia |  | Hyperglycemia |  |
| --- | --- | --- | --- | --- | --- | --- | --- | --- |
|  | HR | p-value | HR | p-value | HR | p-value | HR | p-value |
| Joint Effects |  |  |  |  |  |  |  |  |
| No T2DM or Dementia | 1<br>[reference] | - | 1<br>[reference] | - | 1<br>[reference] | - | 1<br>[reference] | - |
| T2DM only | 1.28 [1.26, 1.3] | <.001*** | 2.02 [1.87, 2.19] | <.001*** | 21.28 [17.76, 25.5] | <.001*** | 11.73 [9.17, 15] | <.001*** |
| Dementia only | 1.89 [1.82, 1.95] | <.001*** | 6.66 [6.14, 7.22] | <.001*** | 14.73 [10.47, 20.71] | <.001*** | 3.39 [1.68, 6.83] | <.001*** |
| Comorbid | 1.9 [1.75, 2.06] | <.001*** | 9.08 [7.57, 10.9] | <.001*** | 104.94 [73.79, 149.24] | <.001*** | 15.94 [6.47, 39.31] | <.001*** |
| Comorbid Effect Modification |  |  |  |  |  |  |  |  |
| Effect of t2dm given dementia | 1.01 [0.92, 1.1] | 0.897 | 1.36 [1.13, 1.65] | 0.002** | 7.13 [4.63, 10.97] | <.001*** | 4.7 [1.55, 14.21] | 0.006** |
| Effect of dementia given t2dm | 1.48 [1.36, 1.61] | <.001*** | 4.49 [3.69, 5.45] | <.001*** | 4.93 [3.6, 6.76] | <.001*** | 1.36 [0.56, 3.31] | 0.509 |
| Interaction |  |  |  |  |  |  |  |  |
| Multiplicative scale | 0.79 [0.72, 0.86] | <.001*** | 0.67 [0.55, 0.83] | <.001*** | 0.33 [0.21, 0.53] | <.001*** | 0.4 [0.13, 1.22] | 0.109 |
| RERI | -0.27 [-0.44, -0.1] | 0.999 | 1.4 [-0.29, 3.09] | 0.052 | 69.93 [35.17, 104.68] | <.001*** | 1.82 [-12.43, 16.07] | 0.401 |
| Covariates |  |  |  |  |  |  |  |  |
| Sex male | 1.14 [1.13, 1.15] | <.001*** | 0.72 [0.69, 0.76] | <.001*** | 1.08 [0.93, 1.25] | 0.336 | 1.11 [0.9, 1.36] | 0.339 |
| Deprivation | 1.06 [1.05, 1.06] | <.001*** | 1.05 [1.04, 1.07] | <.001*** | 1.16 [1.1, 1.22] | <.001*** | 1.09 [1.02, 1.17] | 0.015* |

\* = $p < 0.05$ , \*\* = $p < 0.01$ , \*\*\* = $p < 0.001$

**Table 5:** Mortality Hazard Ratios.

| Measures | HR | p-value |
| --- | --- | --- |
| Joint Effects |  |  |
| No T2DM or Dementia | 1 [reference] | - |
| T2DM-only | 2.58 [2.47, 2.68] | <.001*** |
| Dementia-only | 14.47 [13.9, 15.05] | <.001*** |
| Comorbid | 15.99 [14.47, 17.66] | <.001*** |
| Comorbid Effect Modification |  |  |
| Effect of t2dm given dementia | 1.11 [1, 1.23] | 0.06 |
| Effect of dementia given t2dm | 6.21 [5.59, 6.89] | <.001*** |
| Interaction |  |  |
| Multiplicative scale | 0.43 [0.38, 0.48] | <.001*** |
| RERI | -0.06 [-1.71, 1.59] | 0.527 |
| Covariates |  |  |
| Sex male | 1.61 [1.57, 1.64] | <.001*** |
| Deprivation | 1.11 [1.1, 1.12] | <.001*** |

